# Performance of ChatGPT in Diagnosis of Corneal Eye Diseases

**DOI:** 10.1101/2023.08.25.23294635

**Authors:** Mohammad Delsoz, Yeganeh Madadi, Wuqaas M Munir, Brendan Tamm, Shiva Mehravaran, Mohammad Soleimani, Ali Djalilian, Siamak Yousefi

## Abstract

**Introduction:** Assessing the capabilities of ChatGPT-4.0 and ChatGPT-3.5 for diagnosing corneal eye diseases based on case reports and compare with human experts.

**Methods:** We randomly selected 20 cases of corneal diseases including corneal infections, dystrophies, degenerations, and injuries from a publicly accessible online database from the University of Iowa. We then input the text of each case description into ChatGPT-4.0 and ChatGPT3.5 and asked for a provisional diagnosis. We finally evaluated the responses based on the correct diagnoses then compared with the diagnoses of three cornea specialists (Human experts) and evaluated interobserver agreements.

**Results:** The provisional diagnosis accuracy based on ChatGPT-4.0 was 85% (17 correct out of 20 cases) while the accuracy of ChatGPT-3.5 was 60% (12 correct cases out of 20). The accuracy of three cornea specialists were 100% (20 cases), 90% (18 cases), and 90% (18 cases), respectively. The interobserver agreement between ChatGPT-4.0 and ChatGPT-3.5 was 65% (13 cases) while the interobserver agreement between ChatGPT-4.0 and three cornea specialists were 85% (17 cases), 80% (16 cases), and 75% (15 cases), respectively. However, the interobserver agreement between ChatGPT-3.5 and each of three cornea specialists was 60% (12 cases).

**Conclusions:** The accuracy of ChatGPT-4.0 in diagnosing patients with various corneal conditions was markedly improved than ChatGPT-3.5 and promising for potential clinical integration.

**Key summary points:**

- The aim of this work was to evaluate the performance of ChatGPT-4 and ChatGPT-3.5 for providing the provisional diagnosis of different corneal eye diseases based on case descriptions and compared them with three cornea specialists.
- The accuracy of ChatGPT-4.0 in diagnosing patients with various corneal conditions was significantly better than ChatGPT-3.5 based on the specific cases.
- The interobserver agreement between ChatGPT-4.0 and ChatGPT-3.5 was 65% while the interobserver agreement between ChatGPT-4.0 and three cornea specialists were 85%, 80%, and 75%, respectively.

## INTRODUCTION

The cornea is a clear, non-vascularized tissue that serves as a structural barrier, offering defense against infections to the eye^1^. Corneal eye diseases encompass a diverse range of conditions, including but not limited to corneal infections, dystrophies, degenerations, and injuries^2^. Identifying corneal diseases can be challenging and time-consuming particularly when access to specialized eye care provider is limited ^3,4^. Accurate and timely diagnosis of these conditions is paramount to preserving visual acuity and ensuring optimal patient outcomes.

In recent years, the integration of artificial intelligence (AI) into various medical disciplines has paved the way for innovative approaches to diagnosis and patient care^5^. Ophthalmology, one of the most imaging intensive fields of medicine, has witnessed a significant transformation with the emergence of AI-powered diagnostic tools^6-8^. However, AI applications in the anterior segment parts of the eye including cornea^9-12^ have received less attention compared to the AI applications in posterior segment of the eye including retina. ^13-16^

Among AI tools, ChatGPT, a cutting-edge large language model (LLM) developed by OpenAI (San Francisco, California), has recently received attention, and holds great potential for comprehending clinical expertise and delivering relevant information^17,18^. ChatGPT employs deep learning techniques to generate coherent and contextually relevant text based on user inputs^19^. This AI-driven tool has shown remarkable capabilities in diverse domains since its inception^20-22^, and its use in the field of ophthalmology is highly promising particularly in the landscape of diagnostics^20,23^.

This article explores the capabilities of ChatGPT-4.0 (commercially available version 4.0, updated on March 13, 2023) and ChatGPT-3.5 (publicly available version 3.5, updated on August 3, 2022) in diagnosing corneal eye diseases based on detailed case descriptions and comparing with human experts. Gaining insight into the capacities and limitations of such tools can shape the creation of enhanced systems for supporting the diagnoses in an automated way. This, in turn, may enhance triaging as well as patient care for those with corneal eye diseases and mitigate the demands for specialized ophthalmic services particularly in underserved regions.

## METHODS

### Case Collection

We selected a total of 20 cases with various corneal eye diseases from the openly available database offered by the Department of Ophthalmology and Visual Sciences at the University of Iowa (https://webeye.ophth.uiowa.edu/eyeforum/cases.htm). These 20 cases were selected from over 200 cases which were categorized based on ophthalmic subspecialty. The underlying corneal conditions included corneal infections, dystrophies, degenerations, and injuries including Acanthamoeba keratitis, Acute corneal Hydrops, Atopic Keratoconjunctivitis, Calcific Band Keratopathy, Cogan’s syndrome, Corneal Marginal Ulcer, Cystinosis, Cytarabine induced keratoconjunctivitis, Exposure Keratopathy, Fabry disease, Fuchs’ Endothelial Corneal Dystrophy, Herpes Simplex Viral (HSV) Keratitis, Infectious Crystalline Keratopathy (ICK), Lattice corneal dystrophy type II (Meretoja’s syndrome), Megalocornea, Peripheral Ulcerative Keratitis, Posterior Polymorphous Corneal Dystrophy (PPCD), Pseudophakic Bullous Keratopathy (PBK), Salzmann’s Nodular Degeneration (SND), and Amiodarone-Induced Corneal Deposits (Corneal Verticillata). Details of every case encompassed patient’s demographics, chief complaint, present illness, and major examination findings. Case reports that required specialized exam maneuvers to establish the diagnosis (e.g., Fungal Keratitis-Fusarium) or case reports that are overly obvious (e.g., Chemical Eye Injury) were excluded. Institutional review board (IRB) approval was not required per the direction of our local IRB office as we used a publicly accessible dataset with no patient’s information in this analysis. This study was compliant to the tenets of the Helsinki declaration and ethical aspects was approved by our local research ethics office.

### ChatGPT

ChatGPT represents a derivation of the GPT (Generative Pre-trained Transformer) language model tailored for producing text within conversational settings. Through extensive refinement on substantial conversational datasets, it possesses the capability to produce pertinent and logically connected responses in correspondence with provided input^24^. ChatGPT-4.0 emerged as OpenAI’s most recent language model, embodying a substantial enhancement over its forerunners. GPT-3 was initially trained based on about 176 billion parameters while GPT-4.0 is trained based on approximately 1.75 trillion parameters^25^. GPT-4.0 stands as an advanced multimodal model that leverages diverse data formats to elevate its performance. As such, ChatGPT possesses some level of computer vision-based image interpretation capabilities that are however not yet appropriate for disease diagnosis^24^.

### ChatGPT Diagnosis

We input identical case descriptions into ChatGPT-4.0 and ChatGPT-3.5 and assessed whether the model was able to provide the correct provisional diagnoses. Specifically, we asked: “What is the most likely diagnosis?” (Fig. 1)

**Figure 1.**
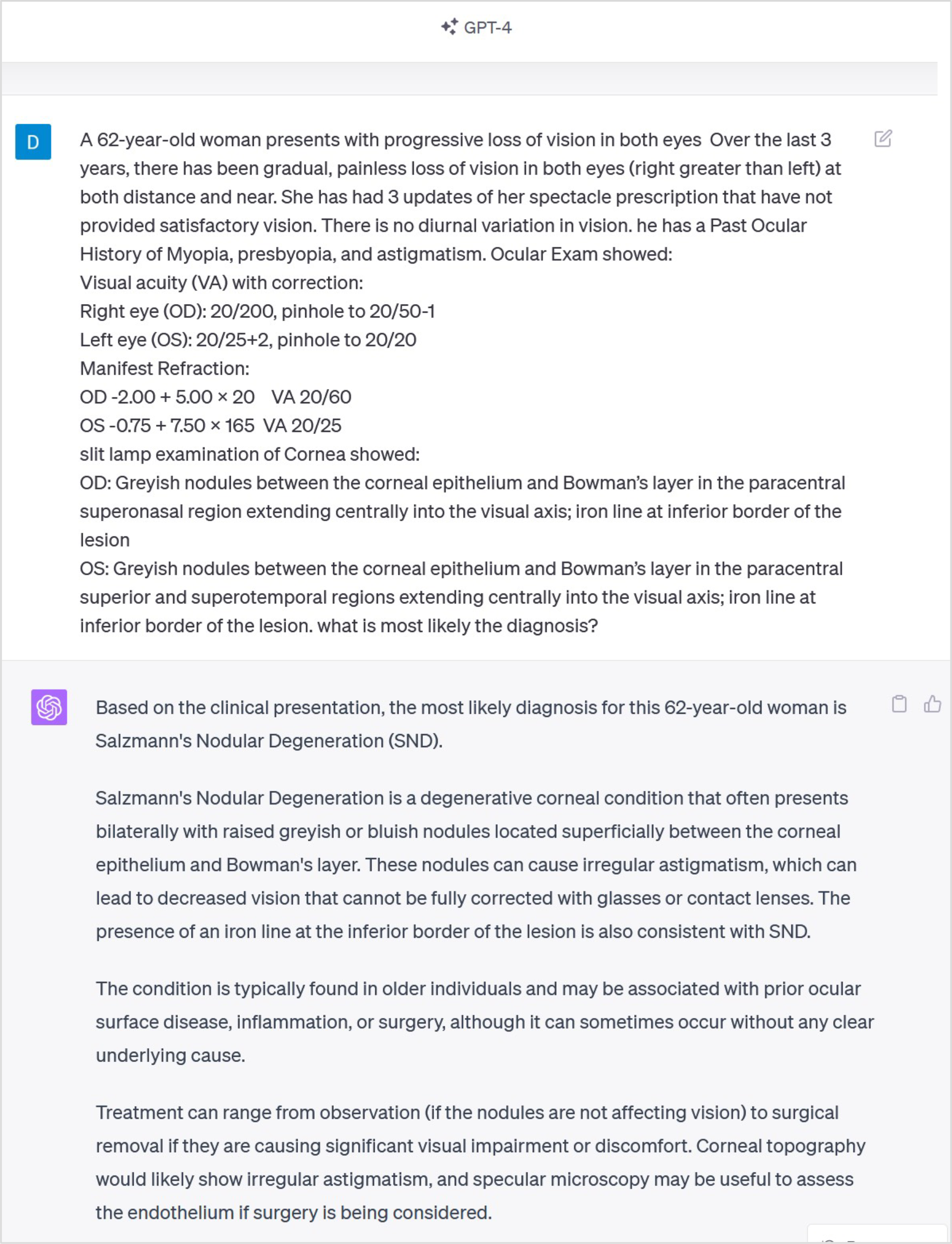
A sample case description input into the ChatGPT-4.0 model and corresponding responses.

We then compared the accuracy of ChatGPT-4.0 and ChatGPT-3.5 with respect to the correct diagnosis. Due to possible reinforcement learning from human feedback (RLHF) capabilities of ChatGPT, we recorded and used the first answer of ChatGPT for the comparisons.

## RESULTS

A comprehensive assessment was conducted, utilizing 20 case presentations spanning various corneal eye diseases, to appraise ChatGPT’s proficiency in generating provisional diagnoses. ChatGPT-4 made the correct diagnosis in 17 out of 20 cases (85%), while ChatGPT-3.5 correctly diagnosed 12 out of 20 cases (60%). Moreover, the three cornea specialists made correct diagnosis of 100% (20 cases), 90% (18 cases), and 90% (18 cases), respectively. The Interobserver agreement between ChatGPT-4.0 and ChatGPT-3.5 was 13 out of 20 cases (65%) while the interobserver agreement between ChatGPT-4.0 and three cornea specialists were 85% (17 cases), 80% (16 cases), and 75% (15 cases), respectively. However, the interobserver agreement between ChatGPT-3.5 and each of three cornea specialists was 60%. Table 1 shows the details of the provisional diagnosis provided by ChatGPT-4.0, ChatGPT-3.5, and human experts. It took approximately 20-40 minutes for the graders to diagnose 20 cases while it took around a couple of minutes (each case around a few seconds) for ChatGPTs to complete diagnosis.

**Table 1.** Provisional diagnoses provided by ChatGPT-4.0, ChatGPT-3.5 and Human Experts.

| No | Corneal Eye diseases | ChatGPT4 Diagnosis | ChatGPT3.5 Diagnosis | Human Expert Diagnosis |
| --- | --- | --- | --- | --- |
| 1 | Cystinosis | Cystinosis | Cystinosis | H1: Cystinosis<br>H2: Cystinosis<br>H3: Cystinosis |
| 2 | Fuchs' Endothelial Corneal Dystrophy (FECD) | FECD | FECD | H1: FECD<br>H2: FECD<br>H3: FECD |
| 3 | Pseudophakic Bullous Keratopathy (PBK) | PBK | **Fuchs' Endothelial Corneal Dystrophy** | H1: PBK<br>H2: PBK<br>H3: PBK |
| 4 | Amiodarone-Induced Corneal Deposits (Corneal Verticillata) | Amiodarone-Induced Corneal Deposits | Amiodarone-Induced Corneal Deposits | H1: Amiodarone-Induced Corneal Deposits<br>H2: Amiodarone-Induced Corneal Deposits<br>H3: Amiodarone-Induced Corneal Deposits |
| 5 | Acanthamoeba Keratitis | Acanthamoeba Keratitis | Acanthamoeba Keratitis | H1: Acanthamoeba Keratitis<br>H2: Acanthamoeba Keratitis<br>H3: Acanthamoeba Keratitis |
| 6 | Cogan's Syndrome (Interstitial Keratitis & Vertigo) | **Scleritis** | **Ocular Rosacea** | H1: Cogan's Syndrome<br>H2: **Episcleritis**<br>H3: Cogan's Syndrome |
| 7 | Infectious Crystalline Keratopathy (ICK) | **Fungal Keratitis** | **Recurrent Herpes Simplex Virus Keratitis** | H1: ICK<br>H2: ICK<br>H3: ICK |
| 8 | Megalocornea | Megalocornea | **Positional Pseudophacodonesis** | H1: Megalocornea<br>H2: Megalocornea<br>H3: Megalocornea |
| 9 | Herpes Simplex Viral Keratitis | Herpes Simplex Viral Keratitis | Herpes Simplex Viral Keratitis | H1: Herpes Simplex Viral Keratitis<br>H2: Herpes Simplex Viral Keratitis<br>H3: Herpes Simplex Viral Keratitis |
| 10 | Atopic Keratoconjunctivitis | Atopic Keratoconjunctivitis | **Ocular Cicatricial Pemphigoid (OCP)** | H1: Atopic Keratoconjunctivitis<br>H2: Atopic Keratoconjunctivitis<br>H3: **OCP** |
| 11 | Lattice Corneal Dystrophy Type II (Meretoja's syndrome) | **Meesmann Corneal Dystrophy (MCD)** | **MCD** | H1: Lattice Corneal Dystrophy Type II<br>H2: Lattice Corneal Dystrophy Type II<br>H3: Lattice Corneal Dystrophy Type II |
| 12 | Salzmann's Nodular Degeneration (SND) | SND | SND | H1: SND<br>H2: SND<br>H3: SND |
| 13 | Exposure Keratopathy | Exposure Keratopathy | Exposure Keratopathy | H1: Exposure Keratopathy<br>H2: Exposure Keratopathy<br>H3: Exposure Keratopathy |
| 14 | Peripheral Ulcerative Keratitis | Peripheral Ulcerative Keratitis | Peripheral Ulcerative Keratitis | H1: Peripheral Ulcerative Keratitis<br>H2: Peripheral Ulcerative Keratitis<br>H3: Peripheral Ulcerative Keratitis |
| 15 | Calcific Band Keratopathy | Calcific Band Keratopathy | **Superficial Corneal Scar** | H1: Calcific Band Keratopathy<br>H2: Calcific Band Keratopathy<br>H3: Calcific Band Keratopathy |
| 16 | Posterior Polymorphous Corneal Dystrophy (PPCD) | PPCD | **Congenital Hereditary Endothelial Dystrophy (CHED)** | H1: PPCD<br>H2: **Granular Corneal Dystrophy**<br>H3: PPCD |
| 17 | Acute Corneal Hydrops | Acute Corneal Hydrops | Acute Corneal Hydrops | H1: Acute Corneal Hydrops<br>H2: Acute Corneal Hydrops<br>H3: Acute Corneal Hydrops |
| 18 | Corneal Marginal Ulcer | Corneal Ulceration | Corneal Ulcer | H1: Corneal Marginal Ulcer<br>H2: Corneal Marginal Ulcer<br>H3: **Mooren Ulcer** |
| 19 | Fabry Disease | Fabry Disease | Fabry Disease | H1: Fabry Disease<br>H2: Fabry Disease<br>H3: Fabry Disease |
| 20 | Cytarabine Induced Keratoconjunctivitis | Cytarabine Induced Keratoconjunctivitis | Cytarabine Induced Keratoconjunctivitis | H1: Cytarabine Induced Keratoconjunctivitis<br>H2: Cytarabine Induced Keratoconjunctivitis<br>H3: Cytarabine Induced Keratoconjunctivitis |

## DISCUSSION

We conducted a prospective study to examine the performance of ChatGPT-4.0 and ChatGPT-3.5 based on 20 cases with different types of corneal eye diseases. The accuracy of ChatGPT-4.0 was 85% while the accuracy of ChatGPT-3.5 was 60%. The interobserver agreement between ChatGPT-4 and the ChatGPT-3.5 was reasonable (65%). We observed that compared to the publicly available ChatGPT-3.5, ChatGPT-4.0, the commercial version, generate markedly improved provisional diagnosis for different corneal eye diseases. These models may assist healthcare providers in generating consistent and useful information regarding the underlying corneal condition.

Some of the capabilities and limitations of ChatGPT in ophthalmology have been discussed previously.^20^ Recently, ChatGPT was investigated in responding to multiple choice questions from the USMLE and it was observed that ChatGPT correctly responded to over 50% of questions and also provided relevant supporting explanations for the selected choices. ^21^ More relevant to our study, a recent investigation showed that ChatGPT-3.0 correctly diagnosed 9 out of 10 general ophthalmology case (90%).^26^ Our accuracy was based on ChatGPT-3.5 was significantly lower (60%) but our ChatGPT-v4.0 accuracy was comparable (85%). Nevertheless, that study assessed general ophthalmology cases while we investigated various corneal conditions that inherently are more challenging to diagnose.

Utilizing conversational AI language models such as ChatGPT could significantly assist frontline healthcare professionals in delivering prompt and precise diagnoses to their patients. In near real-time, such language models could assist primary care and emergency doctors in not only assessing and treating patients but also directing patients to specialized care when required.

Successful integration of ChatGPT or similar LLMs into ophthalmology and cornea services may offer multifaceted benefits. Firstly, ChatGPT’s capability to quickly process large amounts of medical data enhances diagnostic speed and efficiency, leading to quicker patient management and consistent identification of underlying conditions. ChatGPT may transform medical education as well. It could enable students and practitioners to generate interactive and case-based learning materials to foster a deeper understanding of ophthalmic diseases^22^. Another benefit of ChatGPT-based models is versatility in responding to various kinds of questions, rather than just offering diagnosis based on input images or disease-related parameters. Indeed, ChatGPT was not initially planned to respond to diagnostic support, however, its capability in learning from large corpus has provided ChatGPT to even be applicable in narrowed and specialized areas of disease diagnosis.

In addition to diagnostic capabilities and educational purposes, ChatGPT has the potential to be used for patient education as well. For instance, ChatGPT, as a tool with natural language processing (NLP) capabilities, can translate complex medical terms into simple and accessible language leading to enhanced active patient participation. Collectively, these versatile capabilities of ChatGPT makes it a potential tool that may enhance diagnostics, education, and patient engagement.

Although ChatGPT-4.0 was relatively accurate in making a correct diagnosis on most of the cases, we observed that human expert is more accurate on rare cases. For instance, both ChatGPT versions were incorrect on two rare cases including ICK (case # 7, Table 1) and Lattice Corneal Dystrophy Type II (case #11, Table 1), human experts were correct on both cases. As such, the use of ChatGPT in real-world clinical practice should be considered with caution.

While ChatGPT presents remarkable advantages from several aspects, its potential limitations should be acknowledged as well. The accuracy of ChatGPT lies on the quality and diversity of the training data that it has been exposed to until September 2021^27^. Therefore, the model may encounter challenges when faced with rare or emerging corneal conditions that lack representation in its training dataset. Additionally, ChatGPT’s recommendations should always be validated based on clinical evaluations, as its insights may be derived from non-scientific and publicly available knowledge and historical cases. As such, ChatGPT may generate responses that appear fluent and believable however it may contain factual inaccuracies, a phenomenon often termed as hallucination^28^.

Although our study is one of the first investigations of ChatGPT capabilities in diagnosing corneal conditions, it has several limitations as well. First, we have used an online and publicly available dataset to evaluate ChatGPT thus there is a concern that this database has been exposed to ChatGPT previously. To address this concern, we reviewed the years that the cases were added to this online database and noticed that case # 20 in Table 1 has been added in 2023 to this database, that is after September 2021 that the latest ChatGPT training completed and both ChatGPT versions were correct on this case. Additionally, both ChatGPT versions were incorrect on numerous cases that have been added to this database prior to September 2021. Therefore, the likelihood that ChatGPT has seen this online database is slight. Second, we have evaluated ChatGPT based on 20 cases thus follow up studies are warranted to evaluate ChatGPT based on larger number of cases to verify our findings. However, obtaining larger databases with a greater number of case reports is highly challenging and requires larger multi-center and multi-disciplinary collaborations. One major obstacle however is ethical considerations and data privacy issues. The utilization of patient data for diagnostic purposes raises concerns about data security and patient confidentiality. Therefore, rigorous safeguards and compliance with regulatory standards are imperative to ensure responsible and ethical use of ChatGPT in cornea research and clinical practice.

The integration of ChatGPT into the diagnosis of corneal eye diseases marks a significant milestone in the evolution of ophthalmic practice. As AI continues to reshape healthcare, ChatGPT’s potential to enhance diagnostic accuracy, expedite patient care, empower medical education, and stimulate research is evident. While challenges exist, a balanced approach that combines AI-generated insights with clinical expertise holds the key to unlocking the full potential of ChatGPT for the diagnosis of corneal conditions. As we peer into the future, the collaboration between AI and ophthalmology promises to redefine the standards of care and elevate patient outcomes in the realm of corneal eye diseases.

## CONCLUSION

Corneal diseases encompass a diverse variety of conditions that could be challenging to diagnose. We showed that the accuracy of ChatGPT-4.0 in diagnosing patients with various corneal eye diseases is promising and such models may enhance corneal diagnostics. Additionally, ChatGPT may improve patient interaction and experience as well as medical education. A balanced approach that combines AI-generated insights with clinical findings holds the promise to enhance eye care.

## Abbreviations

LLM: Large Language Model
AI: Artificial Intelligence
ChatGPT: Chat Generative Pretrained Transformer
CK: Infectious Crystalline Keratopathy
PPCD: Posterior Polymorphous Corneal Dystrophy
PBK: Pseudophakic Bullous Keratopathy
SND: Salzmann’s Nodular Degeneration
IRB: Institutional Review Board
RLHF: Reinforcement Learning from Human Feedback
NLP: Natural Language Processing
FECD: Fuchs’ Endothelial Corneal Dystrophy
MCD: Meesmann Corneal Dystrophy
CHED: Congenital Hereditary Endothelial Dystrophy

## Author Contribution

Mohammad Delsoz: Research design, data acquisition and research execution, data analysis and interpenetrations, manuscript preparation

Yeganeh Madadi: Research design

Wuqaas M Munir: Data interpretations and manuscript preparation

Brendan Tamm: Data interpretations

Shiva Mehravaran: Research design

Mohammad Soleimani: Data interpretations

Ali Djalilian: Research design

Siamak Yousefi: Research design, data analysis and interpenetrations, manuscript preparation

## Funding

This work was supported by NIH Grants R01EY033005 (SY), R21EY031725 (SY), grants from Research to Prevent Blindness (RPB), New York (SY), and supports from the Hamilton Eye Institute (SY). The funders had no role in study design, data collection and analysis, decision to publish, or preparation of the manuscript.

## Medical Writing/Editorial Assistance

Not applicable.

## Data Availability

Dataset is online and publicly available.

## Ethical Approval

Institutional review board (IRB) approval was not required per the direction of our local IRB office as we used a publicly accessible dataset with no patient’s information in this analysis. This study was compliant to the ethical tenets of the Helsinki declaration and was approved by our local ethical team.

## Conflict of Interest

Mohammad Delsoz: None.

Yeganeh Madadi: None

Wuqaas M Munir: None

Brendan Tamm: None

Shiva Mehravaran: None

Mohammad Soleimani: None

Ali Djalilian: None

Siamak Yousefi: Remidio, M&S Technologies, Visrtucal Fields, InsihgtAEye, Enolink

